# Autoantigen characterization in the lower esophageal sphincter muscle of patients with achalasia

**DOI:** 10.1101/2021.10.01.21264384

**Authors:** Ángel Priego-Ranero, Ghislain Opdenakker, Norma Uribe-Uribe, Diana Aguilar-León, Carlos A. Nuñez-Álvarez, Diego F. Hernández-Ramírez, Elizabeth Olivares-Martínez, Enrique Coss-Adame, Miguel A. Valdovinos, Janette Furuzawa-Carballeda, Gonzalo Torres-Villalobos

**Author notes:** **Correspondence** Gonzalo Torres-Villalobos, Departments of Experimental Surgery and Surgery, Instituto Nacional de Ciencias Médicas y Nutrición Salvador Zubirán, Tlalpan, Mexico CDMX, Mexico and Janette Furuzawa-Carballeda, Department of Immunology and Rheumatology, Instituto Nacional de Ciencias Médicas y Nutrición Salvador Zubirán, Tlalpan, Mexico CDMX, Mexico. Authors share co-first authorship (Ángel Priego-Ranero, Ghislain Opdenakker, Norma Uribe-Uribe, Diana Aguilar-León).

## Abstract

**OBJECTIVE:** To characterize in sera anti-myenteric autoantibody profiles and in tissues MMP-9 proteoforms towards the identification of possible autoantigenic proteins in the muscle of the lower esophageal sphincter (LES) of achalasia patients.

**METHODS:** Biopsies of the LES muscle from 36 achalasia patients, 6 esophagogastric junction outflow obstruction (EGJOO) patients, and 16 transplant donors (TD) were compared in a blind cross-sectional study. Histological characteristics such as inflammation, fibrosis, presence of ganglion cells, cells of Cajal, GAD65, PNMA2, S100, P substance, and MMP-9 proteforms in tissue were assessed by H&E and Picro-Sirius Red stainings, and immunohistochemistry analysis. Antineuronal antibodies (amphiphysin, CV2, and PNMA2 (Ma2/Ta)), onconeural antigens (Ri, Yo, and Hu), recoverin, SOX-1, titin, zic4, GAD65, and Tr (DNER) were evaluated by immunoblot/line assay.

**RESULTS:** Tissue of achalasia patients had heterogeneous inflammatory infiltrates with fibrosis and contrasting higher levels of activated MMP-9 compared with EGJOO and TD. Moreover, lower ganglion cell percentages and cell of Cajal percentages were determined in esophageal tissues of achalasia patients *vs*. TD. In addition, tissue of achalasia patients had higher GAD65 and PNMA2 protein expression *vs*. EGJOO. Unexpectedly, these proteins were absent in TD tissue. S100 and P substance had similar expression levels in tissues of achalasia patients *vs*.TD and EGJOO. Most of the achalasia sera had anti-GAD65 (83%) and anti-PNMA2 (90%) autoantibodies *vs*. EGJOO (17% and 33%, respectively) and healthy volunteers (10% and 0%, respectively).

**CONCLUSION:** Tissue-specific ectopic expression of GAD65 and PNMA/Ta2 and active MMP-9, associated with the presence of specific autoantibodies directed against these proteins, might participate in the pathophysiology of achalasia triggering and/or perpetuating autoimmune disease.

WHAT YOU NEED TO KNOW?

BACKGROUND AND CONTEXT
Achalasia is a primary esophageal motility disorder associated with a selective loss of inhibitory neurons in the myenteric esophageal plexus.
An inflammatory/autoimmune response seems involved in pathophysiology.

NEW FINDINGS
Most of the achalasia sera have anti-GAD65 and anti-PNMA2 autoantibodies vs. EGJOO and healthy volunteers.
Esophageal tissues of achalasia patients have higher ectopic GAD65 and PNMA2 protein expression vs. EGJOO. These proteins are absent in healthy donors with non-inflamed muscle of the LES.
Activated MMP-9 is detected *in situ* together with GAD65 and PNMA2 substrates.

LIMITATIONS
By its design, our study does not shed light on the mechanisms that induce the ectopic expression of the GAD65 and PNMA/Ta2 proteins in LES muscle tissue. It remains impossible to firmly state the sequence of events, e.g. whether the expression of ectopic antigens is due to tissue destruction or the presence of autoantibodies. Whether serum autoantibodies react with autoantigens of the LES muscle tissue of the patients with achalasia also remains unknown.

IMPACT
This information yields new insights into pathophysiological mechanisms of achalasia for better disease understanding and possible design of novel therapeutic strategies.

**LAY SUMMARY:** Most of the achalasia sera have anti-GAD65 and anti-PNMA2. Esophageal tissues of achalasia patients have ectopic GAD65 and PNMA2. This information yields new insights into pathophysiological mechanisms of achalasia for better disease understanding.

---

**I**nflammation is a central mechanism for dealing with insults to tissues, either from pathogenic invaders or by other damage-inducing means. Inflammation is a process to remove the threat, to heal tissues and to return to a state of homeostasis.

However, if an insult persists, it has been hypothesized that inflammation and gradual chronic tissue damage through extracellular proteolytic enzymes, in a context of genetic susceptibility, accompanied by the failure of peripheral tolerance mechanisms, may lead to autoimmunity.^1^ Extracellular proteases may play a crucial role by degrading proteins into remnant fragments, which often constitute immunodominant epitopes. These remnant epitopes are presented to autoreactive T lymphocytes by direct loading into major histocompatibility complex (MHC) molecules or after classical antigen uptake, processing, and MHC presentation. Also, posttranslationally modified remnant peptides may stimulate B cells to produce autoantibodies. This mechanism may act under the condition that active proteases encounter cognate substrates *in situ* and it forms the basis of the “Remnant Epitopes Generate Autoimmunity” (REGA) model.^2^ Presently, it has become a paradigm in which remnant epitopes generate, maintain, and regulate autoimmunity, depending on genetic and epigenetic influences and in a disease phase-specific way. It has been suggested that local proteolysis by matrix metalloproteinases (MMPs) of the myenteric plexus of lower esophageal sphincter (LES) muscle from patients with achalasia can contribute to autoantigen processing into remnant epitopes. In fact, the presence of these enzymes *in situ* in affected esophageal tissues and autoantigen cleavage studies have resulted in the discovery of potential remnant epitopes in achalasia.^1,3^ It has also been proposed that this occurs associated with autoantibodies that target various molecules, including modified self-epitopes of remnant epitopes.^4,5^

In this study, we evaluated potential remnant peptides and autoantibodies in tissue and sera of patients with achalasia and provide evidence that activated MMP-9 and glutamic acid decarboxylase (GAD65) and proteins in the nucleoli of the neuronal cell nuclei, Ma2/Ta (PNMA2) as autoantigens are co-expressed locally in esophageal tissues, specifically in achalasia.

## Materials and Methods

### Patients

This was an exploratory, observational, and cross-sectional study. Thirty-six achalasia patients (type I n=16; type II n=17; type III n=3) and 6 functional esophagogastric junction outflow obstruction (EGJOO) patients were enrolled. The sera and tissue samples were used for histology, immunohistochemistry, and western blot analysis. The samples were obtained between January 2016 and June 2019. All patients and tissue donors were recruited from the Outpatient Clinics of Gastroenterology and Surgery of the Instituto Nacional de Ciencias Médicas y Nutrición Salvador Zubirán (a tertiary referral center in Mexico City, Mexico). Esophagram, HRM (classified based on Chicago v4.0), and upper GI endoscopy were performed to diagnose achalasia and EGJOO.^6,7^ Patients were excluded from the study if they presented any of the following diagnoses: Chagas disease, eosinophilic esophagitis, esophageal stricture, gastric or esophageal cancer, scleroderma, hiatal hernias, gastroesophageal reflux disease with erosive esophagitis, human immunodeficiency virus (HIV), or hepatitis C virus (HCV) infections. None of the patients had been taken opioids. Demographic, clinical, and laboratory information was collected (Table <u>1</u>). As tissue controls, LES muscle samples from multi-organ donors for transplantation (n =16) and EGJOO (n =6) were included. None of the transplant donors had previously known metabolic, inflammatory, neoplastic, or autoimmune diseases. Viral infections with cytomegalovirus (CMV), HCV, hepatitis B virus (HBV), and human immunodeficiency virus (HIV) were excluded. Tests for syphilis and serum ANAs were negative. The cause of death in 9 of the transplant donors was by brain injury, 3 by subarachnoid bleeding, and 4 by hemorrhagic stroke. Demographic, clinical, and laboratory information was collected (Table <u>1</u>).

**Table 1.**
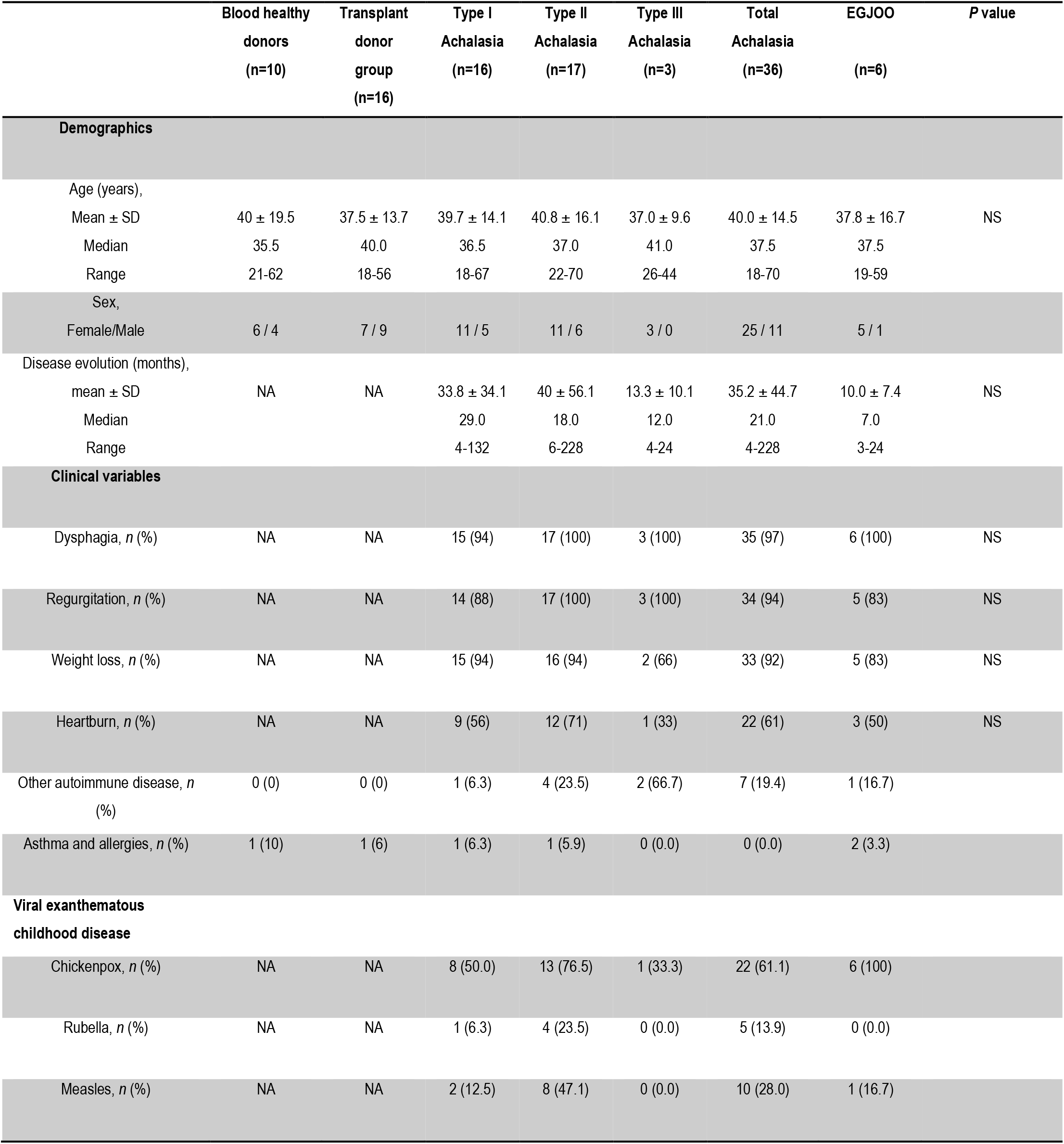
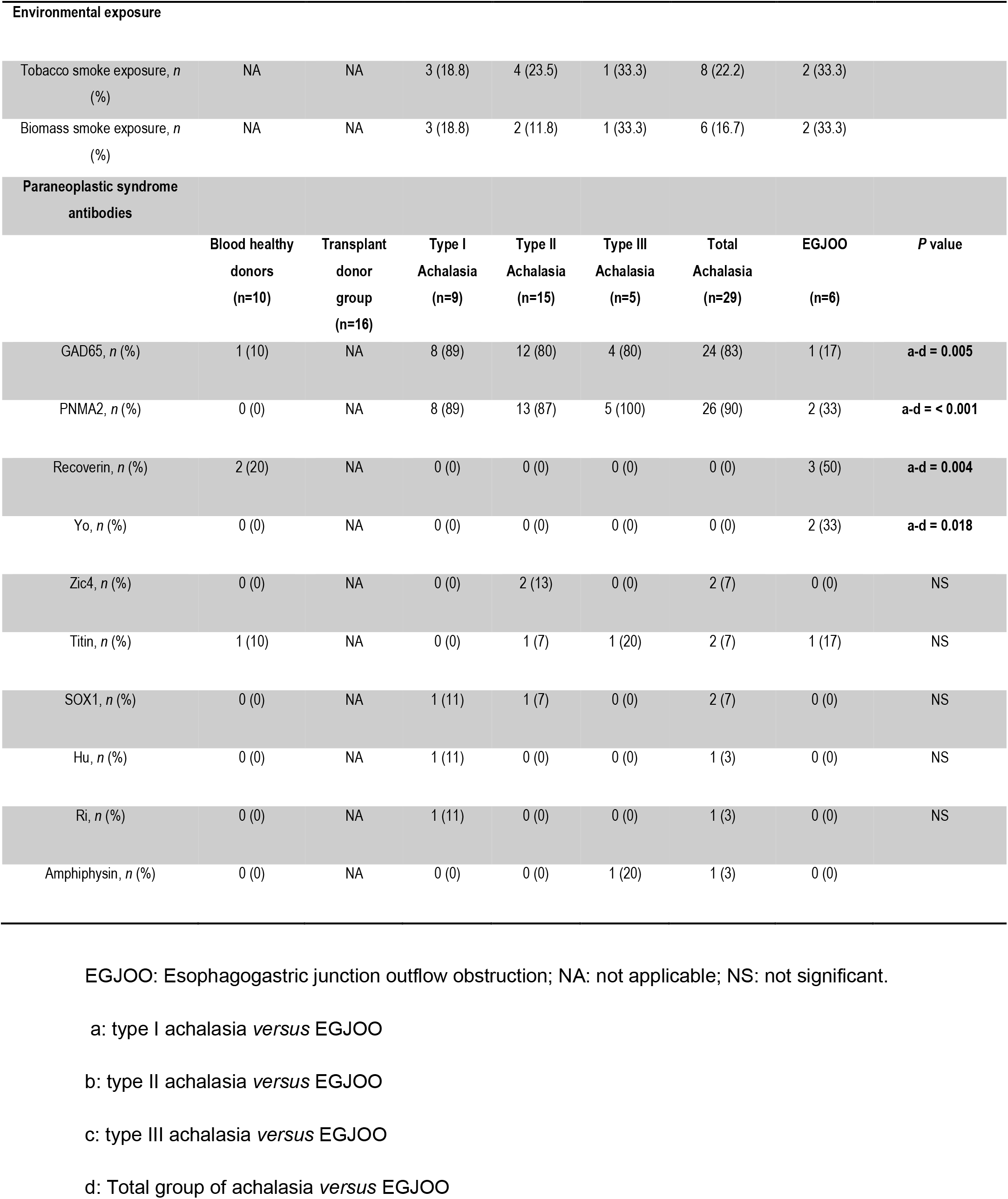
Demographic, clinical, and laboratory variables

|  | Blood healthy<br>donors<br>(n=10) | Transplant<br>donor<br>group<br>(n=16) | Type I<br>Achalasia<br>(n=16) | Type II<br>Achalasia<br>(n=17) | Type III<br>Achalasia<br>(n=3) | Total<br>Achalasia<br>(n=36) | EGJOO<br>(n=6) | P value |
| --- | --- | --- | --- | --- | --- | --- | --- | --- |
| **Demographics** |  |  |  |  |  |  |  |  |
| Age (years), |  |  |  |  |  |  |  |  |
| Mean ± SD | 40 ± 19.5 | 37.5 ± 13.7 | 39.7 ± 14.1 | 40.8 ± 16.1 | 37.0 ± 9.6 | 40.0 ± 14.5 | 37.8 ± 16.7 | NS |
| Median | 35.5 | 40.0 | 36.5 | 37.0 | 41.0 | 37.5 | 37.5 |  |
| Range | 21-62 | 18-56 | 18-67 | 22-70 | 26-44 | 18-70 | 19-59 |  |
| Sex, |  |  |  |  |  |  |  |  |
| Female/Male | 6 / 4 | 7 / 9 | 11 / 5 | 11 / 6 | 3 / 0 | 25 / 11 | 5 / 1 |  |
| Disease evolution (months), |  |  |  |  |  |  |  |  |
| mean ± SD | NA | NA | 33.8 ± 34.1 | 40 ± 56.1 | 13.3 ± 10.1 | 35.2 ± 44.7 | 10.0 ± 7.4 | NS |
| Median |  |  | 29.0 | 18.0 | 12.0 | 21.0 | 7.0 |  |
| Range |  |  | 4-132 | 6-228 | 4-24 | 4-228 | 3-24 |  |
| **Clinical variables** |  |  |  |  |  |  |  |  |
| Dysphagia, *n* (%) | NA | NA | 15 (94) | 17 (100) | 3 (100) | 35 (97) | 6 (100) | NS |
| Regurgitation, *n* (%) | NA | NA | 14 (88) | 17 (100) | 3 (100) | 34 (94) | 5 (83) | NS |
| Weight loss, *n* (%) | NA | NA | 15 (94) | 16 (94) | 2 (66) | 33 (92) | 5 (83) | NS |
| Heartburn, *n* (%) | NA | NA | 9 (56) | 12 (71) | 1 (33) | 22 (61) | 3 (50) | NS |
| Other autoimmune disease, *n* (%) | 0 (0) | 0 (0) | 1 (6.3) | 4 (23.5) | 2 (66.7) | 7 (19.4) | 1 (16.7) |  |
| Asthma and allergies, *n* (%) | 1 (10) | 1 (6) | 1 (6.3) | 1 (5.9) | 0 (0.0) | 0 (0.0) | 2 (3.3) |  |
| **Viral exanthematous**<br>**childhood disease** |  |  |  |  |  |  |  |  |
| Chickenpox, *n* (%) | NA | NA | 8 (50.0) | 13 (76.5) | 1 (33.3) | 22 (61.1) | 6 (100) |  |
| Rubella, *n* (%) | NA | NA | 1 (6.3) | 4 (23.5) | 0 (0.0) | 5 (13.9) | 0 (0.0) |  |
| Measles, *n* (%) | NA | NA | 2 (12.5) | 8 (47.1) | 0 (0.0) | 10 (28.0) | 1 (16.7) |  |

| Environmental exposure |  |  |  |  |  |  |  |  |
| --- | --- | --- | --- | --- | --- | --- | --- | --- |
| Tobacco smoke exposure, *n* (%) | NA | NA | 3 (18.8) | 4 (23.5) | 1 (33.3) | 8 (22.2) | 2 (33.3) |  |
| Biomass smoke exposure, *n* (%) | NA | NA | 3 (18.8) | 2 (11.8) | 1 (33.3) | 6 (16.7) | 2 (33.3) |  |
| Paraneoplastic syndrome antibodies |  |  |  |  |  |  |  |  |
|  | Blood healthy donors (n=10) | Transplant donor group (n=16) | Type I Achalasia (n=9) | Type II Achalasia (n=15) | Type III Achalasia (n=5) | Total Achalasia (n=29) | EGJOO (n=6) | *P* value |
| GAD65, *n* (%) | 1 (10) | NA | 8 (89) | 12 (80) | 4 (80) | 24 (83) | 1 (17) | **a-d = 0.005** |
| PNMA2, *n* (%) | 0 (0) | NA | 8 (89) | 13 (87) | 5 (100) | 26 (90) | 2 (33) | **a-d = < 0.001** |
| Recoverin, *n* (%) | 2 (20) | NA | 0 (0) | 0 (0) | 0 (0) | 0 (0) | 3 (50) | **a-d = 0.004** |
| Yo, *n* (%) | 0 (0) | NA | 0 (0) | 0 (0) | 0 (0) | 0 (0) | 2 (33) | **a-d = 0.018** |
| Zic4, *n* (%) | 0 (0) | NA | 0 (0) | 2 (13) | 0 (0) | 2 (7) | 0 (0) | NS |
| Titin, *n* (%) | 1 (10) | NA | 0 (0) | 1 (7) | 1 (20) | 2 (7) | 1 (17) | NS |
| SOX1, *n* (%) | 0 (0) | NA | 1 (11) | 1 (7) | 0 (0) | 2 (7) | 0 (0) | NS |
| Hu, *n* (%) | 0 (0) | NA | 1 (11) | 0 (0) | 0 (0) | 1 (3) | 0 (0) | NS |
| Ri, *n* (%) | 0 (0) | NA | 1 (11) | 0 (0) | 0 (0) | 1 (3) | 0 (0) | NS |
| Amphiphysin, *n* (%) | 0 (0) | NA | 0 (0) | 0 (0) | 1 (20) | 1 (3) | 0 (0) |  |

EGJOO: Esophagogastric junction outflow obstruction; NA: not applicable; NS: not significant.
a: type I achalasia *versus* EGJOO
b: type II achalasia *versus* EGJOO
c: type III achalasia *versus* EGJOO
d: Total group of achalasia *versus* EGJOO

### High-Resolution Esophageal Manometry (HRM)

An esophageal HRM was performed in every patient at baseline and before being referred for surgery. A solid-state HRM probe with 36 circumferential sensors was used (Medtronic©, Minneapolis, MN, USA). Having the patient in a sitting position and at 45 degrees, stationary esophageal HRM was performed. After a 12 h fasting period, the probe was inserted trans-nasally until passing the esophagogastric junction, assessed visually on the computer screen. Ten water swallows of 5 mL separated by 30 seconds were provided. Analyses were performed using Manoview 2.0 (Medtronic©), and patients were classified according to the latest Chicago classification v4.0^6,7^ into four groups: (a) type I achalasia (without pressurization within the esophageal body), (b) type II (with >20% panpressurization), (c) type III (spastic) and (d) EGJOO (elevated median IRP, ≥20% swallows with elevated intrabolus pressure with evidence of peristalsis). The classification was performed by two gastroenterologists (EC-A, MAV) experts in esophageal HRM.

### HRM Data Analysis

Analysis was performed with ManoView software (Given Imaging, Yokneam, Israel). Four critical metrics of HRM, the distal contractile integral (DCI), the basal EGJ pressure, the intrabolus pressure (IBP), and the integrated relaxation pressure (IRP), were used to assess the pressure motor activity of the esophageal body and EGJ.

Achalasia was defined by IRP >15 mm Hg and aperistalsis. Type I achalasia was defined by absent peristalsis with no compartmentalization of intrabolus pressure. Type II achalasia was defined as panesophageal pressurization in at least 20% swallows in a 30-mmHg isobaric contour. Type III achalasia was characterized by premature contractions with shortened distal latency (<4.5 sec) in at least 20% of swallows. EGJOO was defined as an elevated median IRP in supine and secondary position, and ≥20% swallows with high intrabolus pressure in the supine and sitting position and ≥20% swallows with elevated intrabolus pressure in the supine position, with evidence of peristalsis and in the absence of secondary structural/pharmacological causes.

### Sera samples

Ten mL samples of venous blood were obtained in SST BD Vacutainer tubes and allowed to stand for 20 minutes at room temperature. Subsequently, the blood samples were centrifuged at 400 g for 20 minutes at 4°C. The sera were obtained under sterile conditions, aliquoted, and stored at -70°C until use. They were only thawed once.

### Immunoblot Analysis

To analyze the presence and specific target antigens of 12 circulating anti-myenteric autoantibodies, sera were tested with the EUROLINE paraneoplastic neurologic syndromes 12 Ag (IgG) qualitative kit (Euroimmun AG, Lübeck, Germany). This test is a membrane strip coated with parallel lines of a highly purified combination of neuronal antigens [amphiphysin (antibodies anti amphiphysin a synaptic protein), CV2 (anti-66 kDa protein antibodies), and PNMA2 (proteins in the nucleoli of the neuronal cell nuclei, Ma2/Ta), onconeural antigens (Ri (anti-neuronal nuclear antibodies-2, ANNA-2), Yo (anti-Purkinje cell autoantibodies, PCA-1), and Hu (anti-neuronal nuclear antibodies-1, ANNA-1), recoverin (anti-23kDa and 65 kDa recoverin), SOX-1 (anti-glia nuclear antibodies, AGNA), titin, zic4, GAD65 (antibodies against the enzyme glutamic acid decarboxylase) and Tr (autoantibodies against Tr, a protein in the cytoplasm of cerebellar Purkinje cells, DNER)] separately. After blot strip blocking, sera of patients and positive control (IgG) were incubated at 1/100 for 1 hour at room temperature on a rocking shaker. To detect the bound antibodies, a second incubation was carried out using alkaline phosphatase-labeled anti-human IgG (enzyme conjugating) for 30 minutes at room temperature on a rocking shaker and then revealed with substrate solution (nitroblue tetrazolium chloride/5-Bromo-4-chloro-3-indolyl phosphate) catalyzing a color reaction. For the interpretation, a EUROLine Scan software (Euroimmun) was used.^8^

### Tissue biopsies

Biopsies of LES muscle tissue of achalasia and EGJOO patients were taken during Heller myotomy. After the myotomy was completed without using energy devices, a full-thickness muscle biopsy (2 mm wide and 2 cm long) was obtained by cutting with scissors and was immediately preserved in formalin. LES samples of transplant donors were included as non-inflamed tissue controls. The esophagogastric junction was obtained during organ procuration, with previous signed informed consent from the family. The esophagogastric junction with 3 cm of the esophagus and 2 cm of the stomach was taken. The tissue was transported at 4°C in Bretschneider’s (Custodiol) solution for a period of 4–6 h. Subsequently, a full-thickness biopsy of the esophagus muscle (including the myenteric plexus) was obtained. Tissue was immediately formalin-fixed and paraffin-embedded.

### Histology and morphometric evaluation of interstitial fibrosis

Histological analysis was performed after routine H&E staining on samples from transplant donors and patients with achalasia or EGJOO. The images were captured using Image-Pro Plus v 5.1.1 (Media Cybernetics Inc.) at a magnification of x200 and x600 to evaluate the tissue architecture, the presence of ganglion cells, and to identify the occurrence of inflammatory infiltrates.

To determine interstitial fibrosis (IF), 4-µm sections were stained with Picro-Sirius Red, a specific stain for collagen (red fibers). For the morphological analysis, 3 to 5 evaluations were performed with the Leica QUIPS image and analysis system (Leica Imaging Systems Ltd, Cambridge, UK). Total area and fibrotic area were measured, and the percentage of fibrosis was estimated.

### Immunohistochemistry

Tissue was immediately formalin-fixed and paraffin-embedded. Four *μ*m thick tissue sections were deparaffinized and rehydrated. Then heat-mediated antigen retrieval with citrate buffer pH 6.0 or EDTA buffer pH 8.0 was performed (Supplementary Table 1). Tissues were incubated for 18 h at 4°C with mouse monoclonal or rabbit polyclonal primary antibodies diluted to the manufacturers’ recommendations (Supplementary Table 1) in a humidified chamber. Binding was detected by incubating sections with a biotinylated donkey or goat anti-mouse or rabbit IgG antibody (ABC Staining System; Santa Cruz Biotechnology). Slides were incubated with horseradish peroxidase- (HRP-) streptavidin and 3,3’-diaminobenzidine (DAB) (Sigma-Aldrich) or alkaline phosphatase and aminoethyl carbazole and counterstained with Mayer’s hematoxylin (Lillies’ modification). Negative controls staining were performed with normal human serum diluted 1:100, instead of primary antibody, and the IHC universal negative control reagent specifically designed to work with rabbit, mouse, and goat antibodies (IHC universal negative control reagent, Enzo Life Sciences, Inc., Farmingdale, NY, USA). The reactive blank was incubated with phosphate buffer 3 saline-egg albumin (Sigma-Aldrich) instead of the primary antibody. Control experiments excluded the presence of nonspecific staining or endogenous enzymatic activities. At least two different sections and two fields were examined for each biopsy. Results are expressed as the mean ± standard error of the mean (SEM) of cells quantified by the program Image-Pro Plus version 5·1·1.

For the detection of gelatinase B/MMP-9 proteoforms we used two different monoclonal antibodies against human MMP-9.^9^ REGA-2D9 recognizes the inactive human pro-MMP-9, whereas REGA-3G12 recognizes the activated proteoform lacking the propeptide. Previously we demonstrated that REGA-3G12 neutralizes the activity of gelatinase B/MMP-9 *in vivo*.^10^ Immunoreactivity with REGA-3G12 constitutes a proxy for activated MMP-9 and *in situ* gelatinase B activity and thus needs to be distinguished from the immunoreactivity with REGA-2D9, the latter of which shows the presence of inactive pro-MMP-9.

### Ganglion cells and myenteric interstitial cells of Cajal analysis

Morphometric analysis of ganglion cells (VIP+ cells) and interstitial cells of Cajal (CD117+ or cKit+ cells) in patients and controls was performed in 5 to 10 photographs at x40. Cells were quantified with the program Image-Pro Express v6.3 (Media Cybernetics Inc.). The area of each measurement was 3.1^e5^µm^2^ (0.028 cm^2^). The number of cells was obtained using the formula: [total number of cells/number of measured fields] X 0.028 cm^2^. Results are expressed as the mean ± standard error of the mean (SEM) of cells.

### Statistical Analysis

Descriptive statistics were performed, and categorical variables were compared using *X*^2^ test or Fisher’s exact test; for analysis of continuous variables, Student *t*-test was employed. One-way analysis of variance on ranks by Dunn’s test method was performed for both pairwise comparisons and comparison *versus* a control group. Statistical analysis was done using the IBP SPSS Statistics version 21 IMB (SPSS Inc., Chicago, IL) for Macintosh. Data were expressed as the median, range, and mean ± standard deviation (SD)/standard error of the mean (SEM). The *P* values ≤ 0.05 were considered as significant. This study conforms to the STROBE statement along with references to STROBE and the broader EQUATOR guidelines.

### Ethical Considerations

This work was performed according to the principles expressed in the Declaration of Helsinki. The Research and Bioethics Committee of the Instituto Nacional de Ciencias Medicas y Nutricion Salvador Zubiran approved the study and written informed consent was obtained from all subjects (Ref. No. 1522).

## Results

### Patient characteristics

Thirty-six patients diagnosed with achalasia were included in the study; 25 (69%) were female participants. Patients had a mean age of 40.0 ± 14.5 years and an overall mean disease duration of 35.3 ± 38.9 (range: 1-140 months). A matched control group of 6 patients with EGJOO was included, with 5 (83%) female participants, with a mean age of 37.8 ± 16.7 years and an overall mean disease duration of 10.0 ± 7.4 (range: 3-24 months). The transplant donor group was also predominantly comprised of women (78%), with a mean age of 37.5 ± 13.7 years (Table 1). Clinical manifestations in patients with achalasia included dysphagia (97%), regurgitation (94%), heartburn (61%), and weight loss (92%). The prevalence of autoimmune comorbidity was 19.4% in achalasia patients (Table 1).

### Detection of autoantibodies that recognize antigens in neurologic tissue

To determine the presence of a family of autoantibodies that recognize antigens in neurologic tissue and which of these are associated with a variety of neurologic manifestations occurring as a result of a disorder, the sera from patients with achalasia were analyzed in strips with a combination of neuronal antigens [amphiphysin, CV2, PNMA2, onconeural antigens (Ri, Yo, and Hu), recoverin, SOX-1, titin, zic4, GAD65, and Tr] separately. The specificity of the antineuronal autoantibodies showed that anti-PNMA/Ta2 and anti-GAD65 antibodies were detected in 90% and 83% of sera from achalasia patients, while in 33% and 17% of sera from EGJOO patients and in 0 and 10% of healthy volunteers (Table 1).

### Histological findings

The myenteric plexus was located between the longitudinal and circular layers of muscularis externa of the esophagus. Transplant organ donor tissues (controls) had a myenteric plexus with normal characteristics and typical morphology of ganglion cells (Figure 1A *upper and lower left panel*). Whereas tissues of patients with achalasia showed abundant and heterogeneous inflammatory infiltrates predominantly enriched of lymphocytes and scant polymorphonuclear leukocytes (plexitis), ganglion cells were almost absent (Figure 1B *upper and lower middle panel*). Interestingly, EGJOO tissues displayed axonal hypertrophy, absence of inflammatory infiltrates and scarce ganglion cells (Figure 1C *upper and lower right panel*).

**Figure 1.**
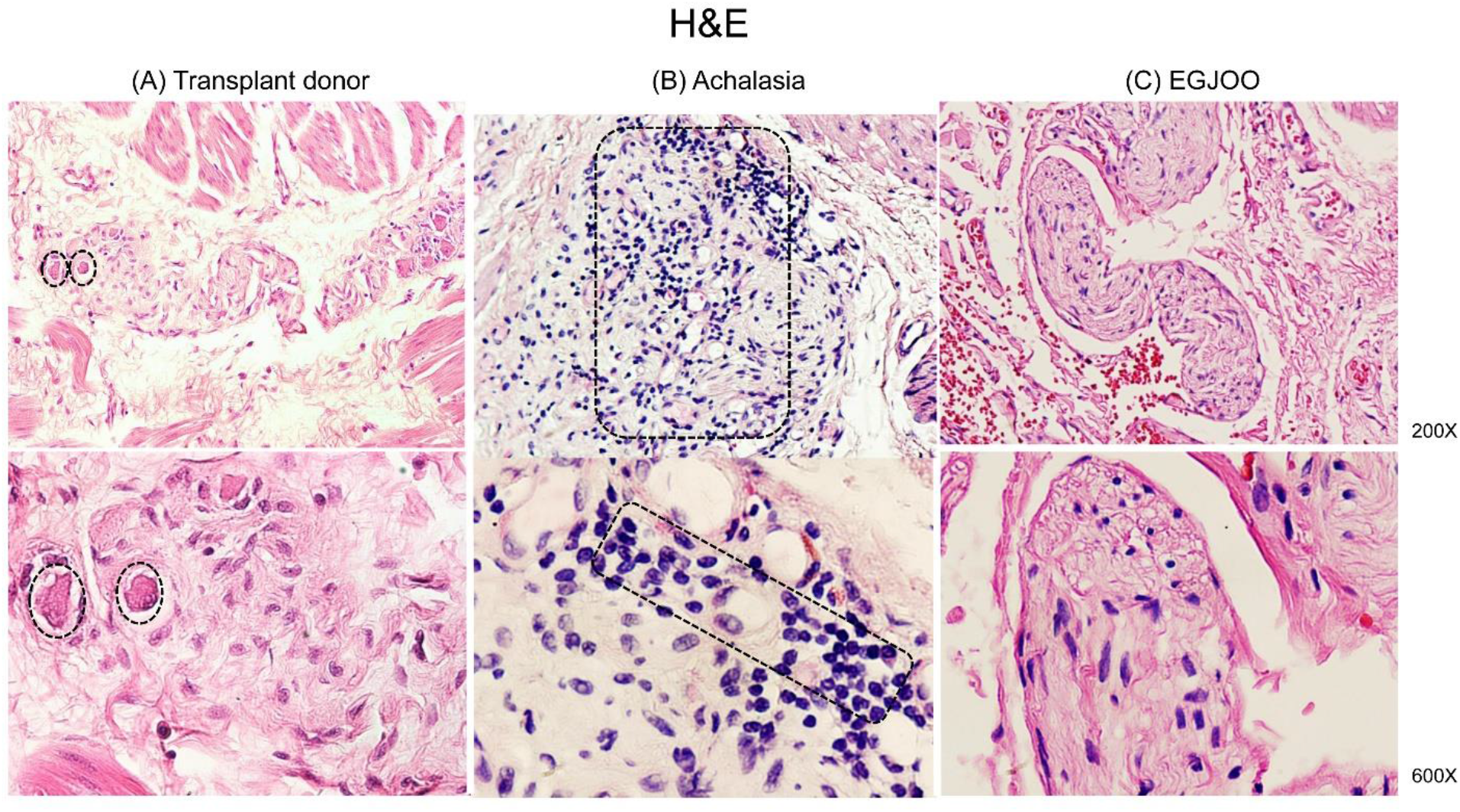
Tissue architecture of myenteric plexus (H&E staining). (*Column* A) Lower esophageal sphincter from transplant donors (n=16). Circles depict ganglion cells. (*Column* B) Lower esophageal sphincter from patients with achalasia (n=36). Ganglion cells are practically absent. Rectangles show heterogeneous inflammatory infiltrate. (*Column* C) Muscle tissue of the lower esophageal sphincter from patients with esophagogastric junction outflow obstruction (EGJOO, n=6). Ganglion cells and inflammatory infiltrates are almost absent. *Upper panel*: magnification x200. *Lower panel*: magnification x600.

### Fibrosis and extracellular matrix turnover proteins in the tissue of patients with achalasia

Regarding fibrosis, transplant organ donors and EGJOO tissues had a normal distribution of collagen fibers (2.3±0.3 and 2.2±0.4%; Figure 2A, C, and D). Nonetheless, the tissue of achalasia patients had a significant increment of dense bundles of collagen fiber depositions (20.7±1.5%, P≤0.001; Figure 2B and D, dark pink). Type I achalasia had the highest level of fibrosis compared with type II and type III achalasia. Besides, MMP-9, an enzyme involved in extracellular matrix degradation in physiological and pathological processes, was determined in the myenteric plexus and blood vessels of esophageal tissue from transplant donor, achalasia, and EGJOO patients, showing a statistically significant increase only in achalasia tissue compared with transplant organ donor and EGJOO tissues (15.1±1.9 *vs*. 3.5±0.5 *vs*. 2.1±0.8; P≤0.001; Figure 3). Most remarkable are the data with the activity-neutralizing monoclonal antibody REGA-3G12, recognizing the active form of MMP-9. Active MMP-9 was clearly increased in achalasia patient biopsies vs those from transplant donors and patients with EGJOO, in particular in the comparisons of the subcohort of type I, II and III achalasia patient samples.

**Figure 2.**
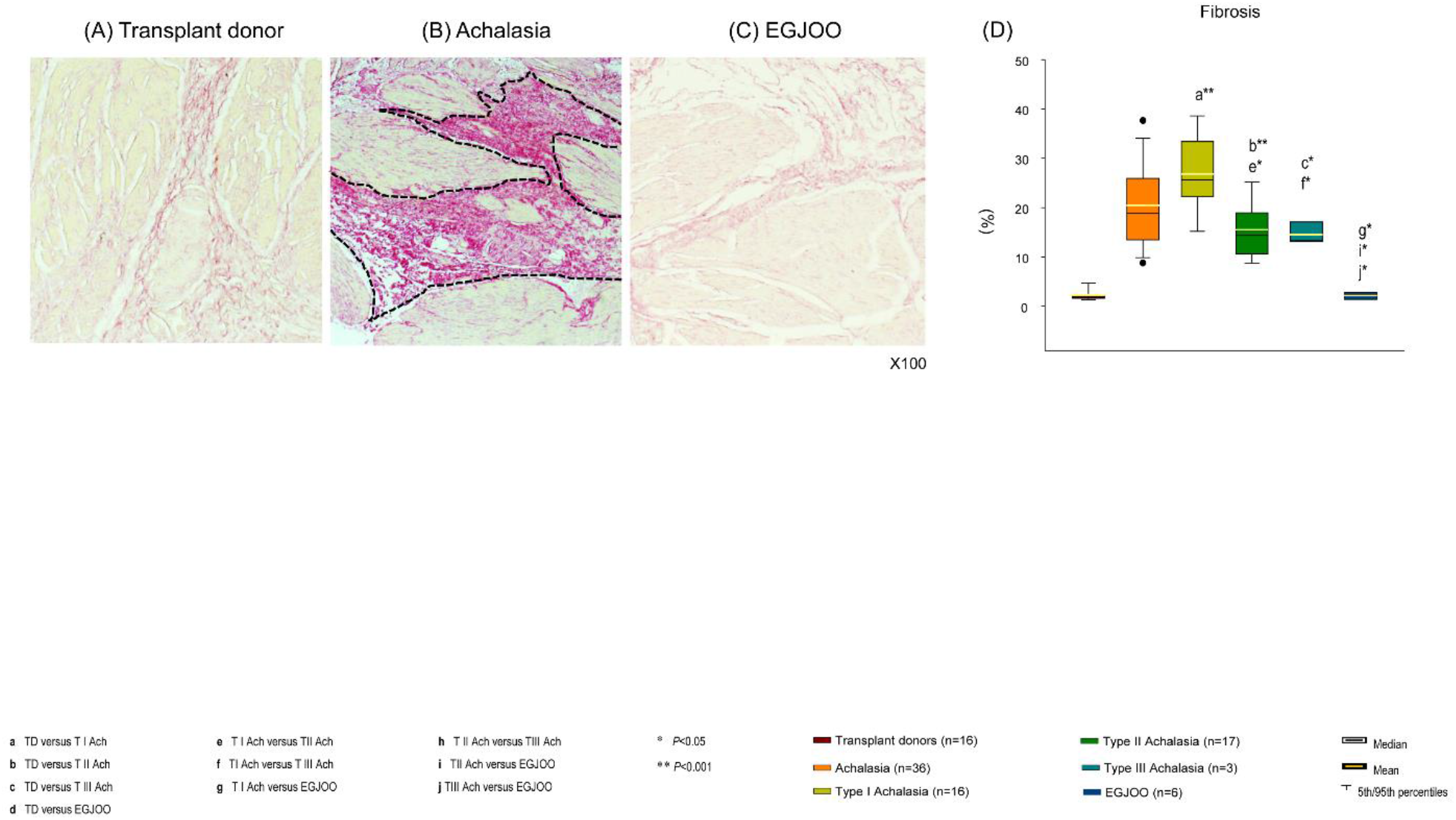
Fibrosis. Representative photomicrograph of interstitial fibrosis in muscle tissues from lower esophageal sphincter of (*column A*) transplant organ donor (n=16), (*column B*) patient with achalasia (n=36), and (*column C*) patient with esophagogastric junction outflow obstruction (EGJOO; *n=6*) stained with Picro-Sirius Red. The area inside the dotted line represents the fibrous tissue (dark pink). Original magnification was x100. (*panel* D) Percentage of fibrosis. The results are expressed as mean (horizontal yellow line), median (horizontal black line), and 5th/95th percentiles. TD: transplant donor; Ach: achalasia; T I Ach: Type I achalasia; T II Ach: Type II achalasia; T III Ach: Type III achalasia. \**P*<0.005; \*\**P*<0.001.

**Figure 3.**
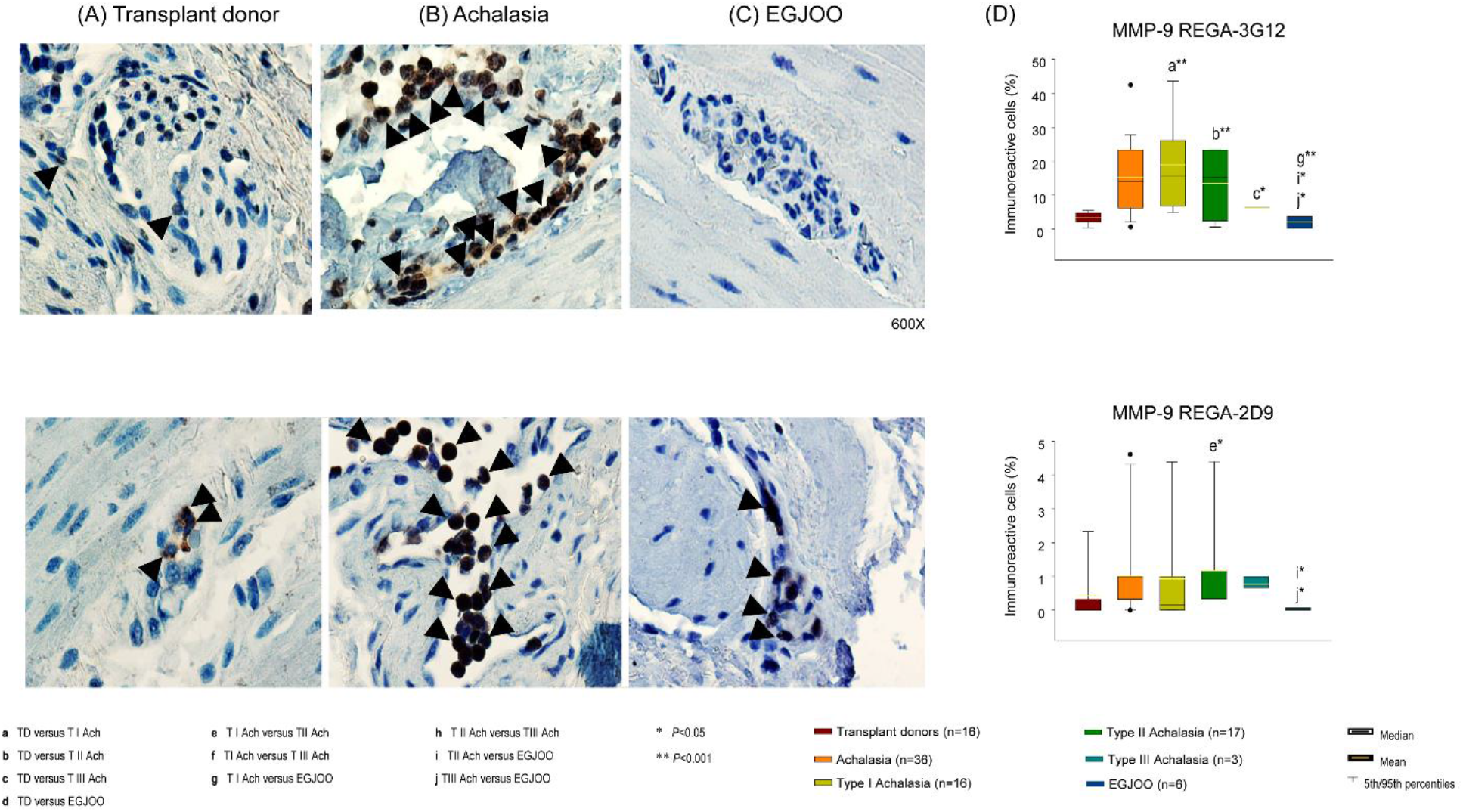
Extracellular matrix turnover. (*Column* A) Representative photomicrograph of MMP-9 in muscle tissue from lower esophageal sphincter of a transplant organ donor (*left panel*); (*Column B*) a patient with achalasia (*middle panel*), and (*Column C*) a patient with esophagogastric junction outflow obstruction (EGJOO; *right panel*). (*Column D*) Percentage of MMP-9 positive cells in tissue biopsies from donors (patients for organ donation, n=16;), achalasia patients (type I, n=16; type II, n=17; and type III, n=3) and EGJOO (n=6). Arrowheads show immunoreactive cells. Original magnification was x600. The upper panel set of photomicrographs and histograms demonstrate the immunoreactivities with REGA-3G12, which is a proxy of *in situ* enzyme activities. The lower panel depicts the immunoreactivity levels with REGA-3G12, yielding data of the presence of inactive proforms of MMP-9. The results are expressed as mean (horizontal yellow line), median (horizontal black line), and 5th/95th percentiles of positive immunoreactive cells. TD: transplant donor; Ach: achalasia; T I Ach: Type I achalasia; T II Ach: Type II achalasia; T III Ach: Type III achalasia. \**P*<0.005; \*\**P*<0.001.

### Ganglion cells and myenteric interstitial cells of Cajal in the tissue of patients with achalasia

Myenteric plexus from achalasia and EGJOO patients had a lower number of ganglion cells (1.3±0.5 and 1.3±1.3 cells/cm^2^) when compared with transplant organ donors (60.2±5.2 cells/cm^2^; P<0.001; Figure 4A and B).

**Figure 4.**
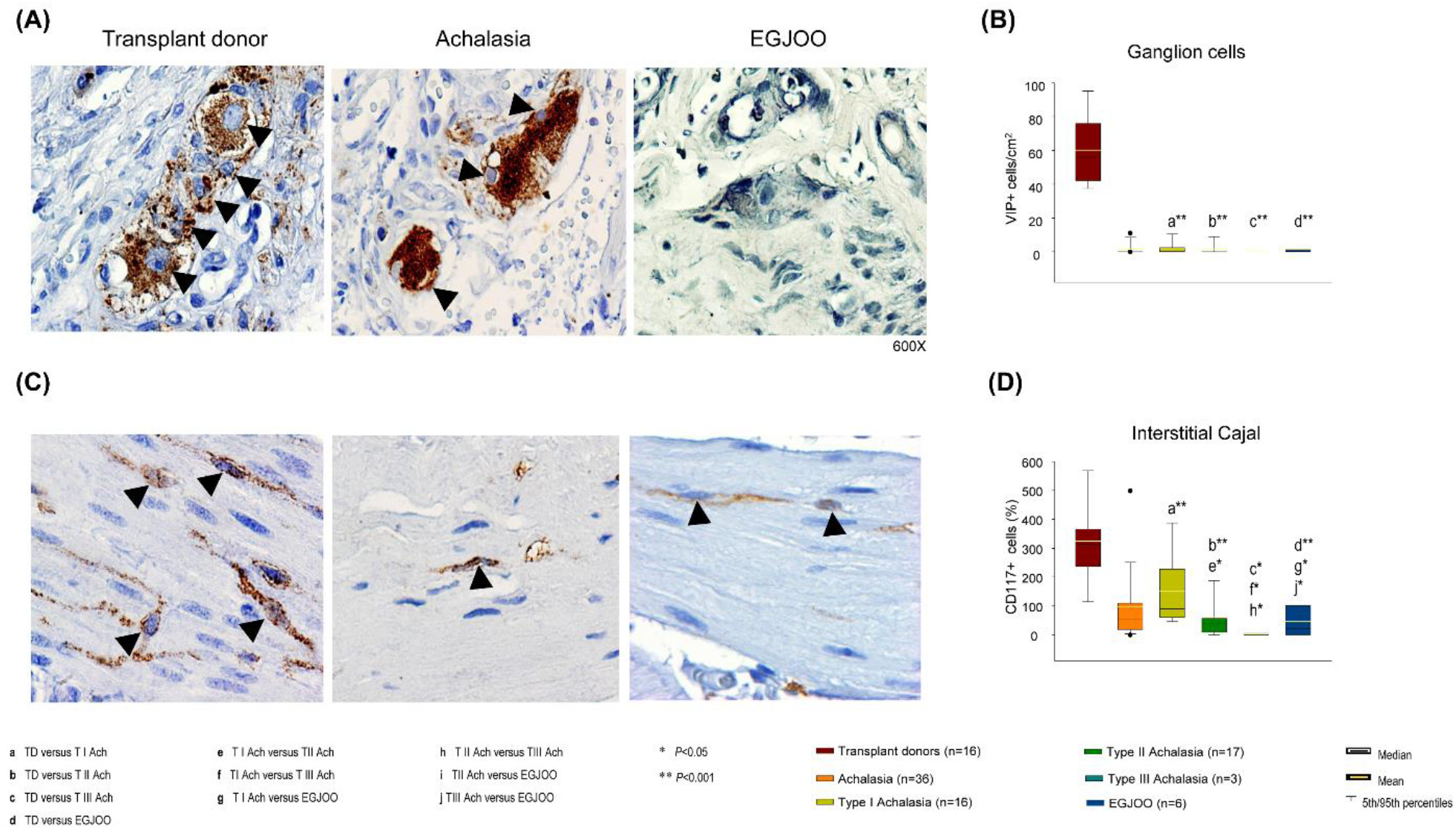
(A) Representative immunostaining of VIP−expressing cells and (C) myenteric interstitial cells of Cajal CD117 (cKit) positive cells in tissue biopsies from transplant organ donors (n=16), achalasia patients (type I, n=16; type II, n=17; and type III, n=3) and esophagogastric junction outflow obstruction (EGJOO, n=6). Arrowheads show immunoreactive cells. Original magnification was x600. (*panel* B, D) Percentage of immunoreactive cells per microscopic field. The results are expressed as mean (horizontal yellow line), median (horizontal black line), and 5th/95th percentiles of positive immunoreactive cells/cm^2^. TD: transplant donor; Ach: achalasia; T I Ach: Type I achalasia; T II Ach: Type II achalasia; T III Ach: Type III achalasia. \**P*<0.005; \*\**P*<0.001.

Moreover, a similar pattern was observed in muscle LES with myenteric interstitial cells of Cajal (transplant organ donors *vs*. achalasia and EGJOO: 337.7±35.9 *vs*. 97.0±22.1 and 49.1±26.2 cells/cm^2^; P<0.001; Figure 4C and D). No statistically significant differences were found between samples from achalasia and EGJOO patients.

### GAD65 (Glutamate decarboxylase 2) and Paraneoplastic antigen Ma2 (onconeural antigen Ma2, PNMA/Ta2 or PNMA2) neuronal proteins detection in the tissue of patients with achalasia

Muscle tissues of the LES of transplant organ donors did not express the GAD65 or PNMA/Ta2 proteins. However, the biopsies of the LES of patients with achalasia type I, II, and III had a tissue-specific ectopic expression of GAD65 at statistically significant levels when compared to transplant organ donors (P<0.001; Figure 5A and B). The patients with type III achalasia had the highest levels of GAD65 compared with EGJOO and transplant organ donors (P<0.001; Figure 5A and B). GAD65 expression in EGJOO patients was also statistically significantly different compared to transplant donors (P<0.001; Figure 5A and B). The PNMA/Ta2 had higher expression levels in patients with achalasia type I, II, and III than transplant organ donors (P<0.001; Figure 5C and D). The patients with type III achalasia had the highest levels of PNMA/Ta2 compared with EGJOO (P<0.024; Figure 5C and D).

**Figure 5.**
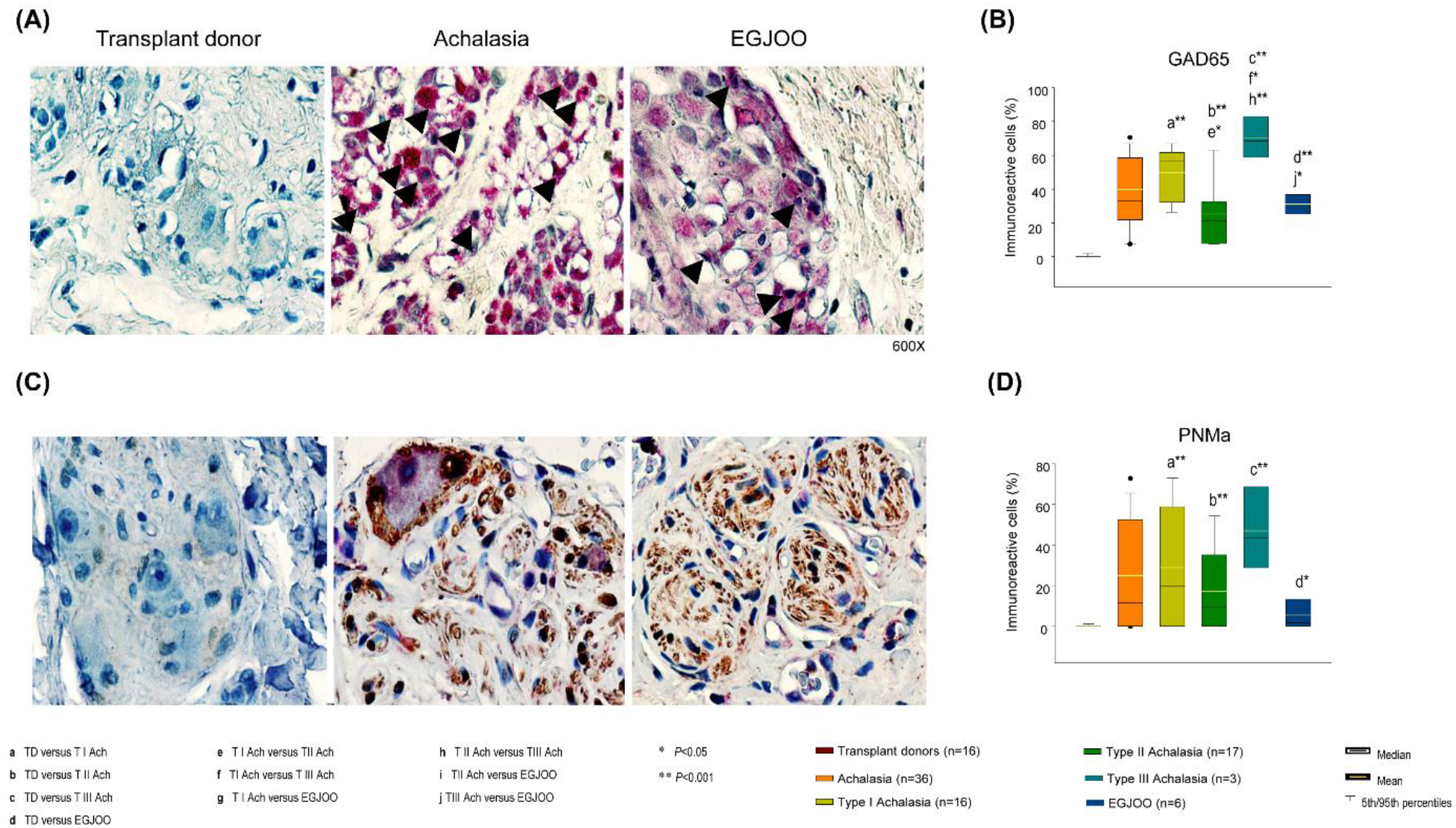
(A) Representative immunostaining of GAD65−expressing cells and (C) PNMa/Ta2 positive cells in tissue biopsies from transplant organ donors (n=16), achalasia patients (type I, n=16; type II, n=17; and type III, n=3) and esophagogastric junction outflow obstruction (EGJOO, n=6). Arrowheads show immunoreactive cells. Original magnification was x600. (*panel* B, D) Percentage of immunoreactive cells per microscopic field. The results are expressed as mean (horizontal yellow line), median (horizontal black line), and 5th/95th percentiles of positive immunoreactive cells/cm^2^. TD: transplant donor; Ach: achalasia; T I Ach: Type I achalasia; T II Ach: Type II achalasia; T III Ach: Type III achalasia. \**P*<0.005; \*\**P*<0.001.

### S100 protein and substance P detection in the tissue of patients with achalasia

Neuronal structures as markers of motility were stained with antibodies against neurofilament, S-100 protein and substance P. The levels of these three antigens were similar in non-subtyped achalasia patients compared with transplant donors (Figure 6A-D), except for substance P, which was found to be higher in the biopsies of patients with type I achalasia vs. the transplant organ donors (P<0.05).

**Figure 6.**
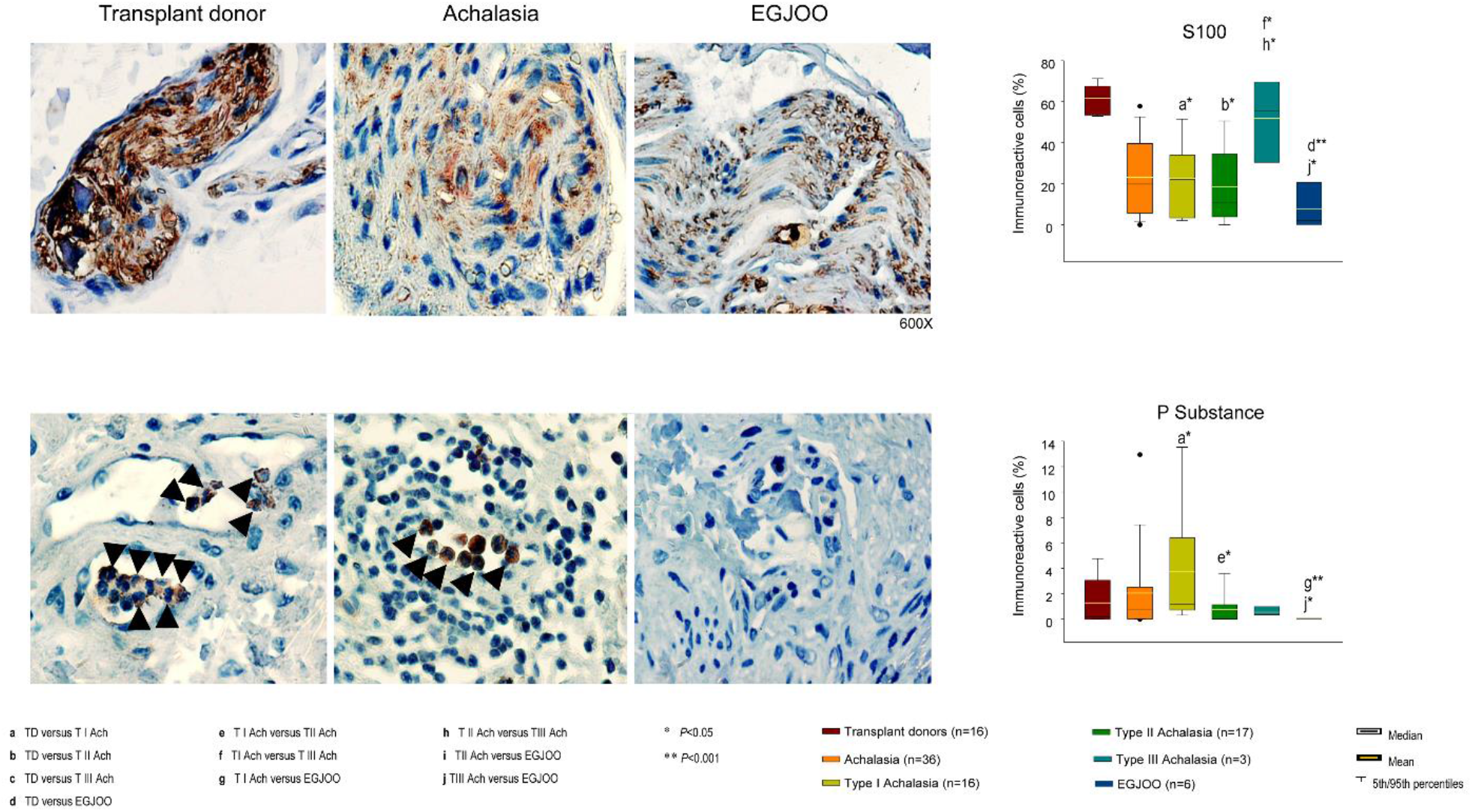
(A) Representative immunostaining of S100−expressing cells and (C) P Substance positive cells in tissue biopsies from transplant organ donors (n=16), achalasia patients (type I, n=16; type II, n=17; and type III, n=3) and esophagogastric junction outflow obstruction (EGJOO, n=6). Arrowheads show immunoreactive cells. Original magnification was x600. (*panel* B, D) Percentage of immunoreactive cells per microscopic field. The results are expressed as mean (horizontal yellow line), median (horizontal black line), and 5th/95th percentiles of positive immunoreactive cells/cm^2^. TD: transplant donor; Ach: achalasia; T I Ach: Type I achalasia; T II Ach: Type II achalasia; T III Ach: Type III achalasia. \**P*<0.005; \*\**P*<0.001.

## Discussion

New concepts have been emerging regarding the hypothesis that achalasia is an autoimmune disease. A viral infection, chronic inflammatory infiltrates, predominantly enriched in lymphocytes that damage the ganglion cells of the Auerbach plexus, the presence in patient sera of organ-specific autoantibodies, the association in the same patient of well-established autoimmune diseases that often occur in the association with one another, either within single individuals or families and genetic factors and polymorphisms, all of these are elements that relate to autoimmune disease.^11^

In this study, we corroborated a number of histopathological hallmarks of achalasia, namely the abundant inflammatory infiltrates predominantly enriched of lymphocytes, accompanied by extensive areas of fibrosis (21% of the tissue), degeneration of the myenteric plexus, plexitis, and more than 97% of loss of ganglion cells, contrasting with the transplant donor group without any sings of tissue inflammation and with histologically normal myenteric plexus.

Extracellular matrix turnover was evaluated through the presence of two MMP-9/gelatinase B proteoforms. The inactive human MMP-9 proform of 92 kDa is reactive with REGA-2D9 and the activated 85 kDa form reacts with REGA-3G12. In previous reports, we showed that the circulating and tissue levels of activated MMP-9 proteoforms are increased in patients with achalasia compared with controls. Moreover, MMP-9 may contribute to the extracellular proteolysis of PNMA/Ta2, Ri, GAD65, and VIP (novel MMP-9 substrates) that might play a role in the pathogenesis of the organ-specific autoimmune disease, including achalasia.^3^ Here we provide the first evidences of *in situ* co-expression of activated MMP-9, a proxy for enzyme activity, and the substrates GAD65 and PNMA/Ta2, the latter of which were defined as achalasia autoantigens by the presence of autoantibodies in individual achalasia patients.

Organ-specific autoimmune diseases occur when a specific adaptive immune response is mounted against self-antigens, causing severe tissue damage. Paul Ehrlich termed this *horror autotoxicus*. Significant progress has been made in defining the targeted molecules in autoimmune diseases, mainly using high titer autoantibodies as probes.^12^ The resulting list of autoantigens, together with careful clinical descriptions of the distinct disease features associated with such responses, has provided a critical framework to understand the mechanisms responsible for antigen selection in autoimmune diseases.^13^

It is noteworthy that the antigenic targets of the highly driven immune responses in autoimmune diseases are limited, quite specific, and often tissue-restricted (e.g., islet cell autoantigens in insulin-dependent diabetes mellitus,^14^ or components of the acetylcholine receptor in myasthenia gravis.^15^

This specificity, while not absolute, has nevertheless allowed autoantibody responses to become clinically valuable markers and/or predictors of phenotype. The mechanisms underpinning this striking specificity remain unclear for many autoantigens. However, accumulating data suggests that properties of the antigens themselves (such as structure, adjuvant properties, and multimeric nature) play central roles in antigen selection.^16^

Two distinct phases in the development of autoimmune diseases are critical: the initiation phase, in which - in many cases - the presence of autoantibodies precedes clinical manifestations, and the propagation phase, in which enhanced autoantigen expression in the target tissue or immune effector pathway-induced antigen generation play essential roles.^1,17^ Some autoantigens, accessed by autoantibodies, are recognized predominantly/exclusively in their native forms. Autoreactive T cells recognize autoantigenic peptides, which are assumed to be biologically relevant during the natural processing of whole protein antigens by intracellular proteasomal or lysosomal proteolysis.^16^

In this vein, autoantibodies against neuronal proteins were detected by a highly analytical and qualitative method identifying IgG class immunoglobulins. In 90% and 83% of the sera from patients with achalasia, anti-PNMA/Ta2 and anti-GAD65 autoantibodies were detected. In comparison, anti-PNMA/Ta2 and anti-GAD65 autoantibodies were found in only 33% and 17% of the sera from patients with EGJOO and in 0% and 10% healthy volunteers. Although with this study we cannot confirm that the detected autoantibodies are pathogenic, the conspicuous ectopic expression of their antigens, PNMA/Ta2, and GAD65, in the muscle tissue of the LES suggests the possibility of a physiopathogenic role for these antibodies. Regarding tissue protein expression levels, it is essential to note that, although RNA-seq studies have been determined that PNMA/Ta2 and GAD65 RNA are present in some squamous epithelial cells, smooth muscle cells, and skeletal muscle cells of the esophagus, in healthy individuals, tissue proteins have not been identified.^18,19^

So far, it is unknown which soluble and cellular factors induce the ectopic expression of GAD65. This expression might be related to at least two different functions, the first one being intensification of synaptic activity, and as the second one, downregulating neuropathic pain through the increased expression of GAD65 to normalized GABA transmission, and ultimately resulting in less sensitivity to pain.^20,21^ However, once the proteins become ectopically expressed, this may be one factor to trigger an autoimmune response.^16,20-23^

PNMA/Ta2 is expressed in the human adult brain, as demonstrated by Northern blot analysis and immunohistochemistry.^24^ It has anti-apoptotic functions. The ectopically PNMA/Ta2 protein accumulation and the presence of MA2 autoantibodies strongly suggest an autoimmune response.^25,26^

In contrast with a previous report by Gockel I et al., in our study, no differences were found in the total achalasia cohort between the presence of intramural nerve cell plexus structures as markers of motility stained with antibodies against neurofilament, S-100 protein, and substance P.^27^ However, type I achalasia patients had higher levels of S100 protein and substance P in comparison with the transplant donors.

We recognize that our study has some limitations, such as this is a cross-sectional study. By its design, it does not shed light on the mechanisms that induce the ectopic expression of the GAD65 and PNMA/Ta2 proteins in the muscle tissue of the LES. It cannot be established which comes first; e.g. whether the expression of ectopic antigens is due to tissue destruction or the presence of autoantibodies. Whether serum autoantibodies react with autoantigens of the LES muscle of the patients with achalasia remains unknown. However, the results suggest a physiopathogenic role for these autoantibodies since the antigens are abundantly expressed in the tissue of patients with achalasia, but not in healthy tissues. Although low expression levels of GAD65 and PNMA/Ta2 were observed in patients with EGJOO, the presence of the respective autoantibodies was not detected.

In addition, our study provided the proof that activated MMP-9 was present *in situ* in achalasia tissue and thus might generate remnant epitopes of autoantigenic proteins. We provided further circumstantial evidence reinforcing the Rega paradigm.^1^ This study contains the first comparison of immunohistochemical analysis of inactive pro-MMP-9, detected by REGA-2D9, and of activated human MMP-9 as determined with the neutralizing monoclonal antibody REGA-3G12.^9,10^ We thus demonstrate that active enzyme and specific substrates come together locally in achalasia. By extracellular proteolysis of autoantigenic proteins *in situ* in achalasia, e.g., by MMP-9, the autoantigenic burden for activation of autoreactive T lymphocytes to provide helper functions for autoantibody formation thus may be increased.

In conclusion, to our knowledge, this is the first study to characterize the presence of the ectopic antigens GAD65 and PNMA/Ta2 in the LES muscle tissue of patients with achalasia and the presence of the respective autoantibodies in their sera. We suggest that both proteins, GAD65 and PNMA2, both being cleaved by MMP-9 *in vitro*^3^ into remnant epitopes define autoantigens that trigger achalasia as an autoimmune disease.

## Data Availability

Data Sharing Statement
Furuzawa. Polymerized type I collagen for COVID-19 treatment. 
Data available: Yes
Data types: Deidentified participant data
How to access data:
When available: beginning date: 01-01-2022
Supporting Documents Document types: None Additional Information Who can access the data: researchers whose proposed use of the data has been approved
Types of analyses: for pre-planned analyses and meta-analyses (if applicable)
Mechanisms of data availability: after approval of a proposal
Any additional restrictions: n/a

## Abbreviations

CMV: cytomegalovirus
DAB: 3,3’-diaminobenzidine
EGJOO: esophagogastric junction outflow obstruction
GAD65: glutamic acid decarboxylase of 65 kDa
HBV: hepatitis B virus
HCV: hepatitis C virus
HIV: human immunodeficiency virus
HRM: High-Resolution (Esophageal) Manometry
HRP: horseradish peroxidase
LES: lower esophageal sphincter
MHC: major histocompatibility complex
MMPs: matrix metalloproteinases
PNMA2: proteins in the nucleoli of the neuronal cell nuclei, Ma2/Ta
REGA: Remnant Epitopes Generate Autoimmunity
SEM: standard error of the mean
TD: transplant donors

## GUARANTOR OF THE ARTICLE

Gonzalo Torres-Villalobos, MD, PhD.

## SPECIFIC AUTHOR CONTRIBUTIONS

A.P.-R., G.O., N.U.-U., D.A.-L., Conceptualization, Materials, Data curation, Formal analysis, Investigation, Methodology, Writing – original draft, Writing – review & editing, C.A.N-Á., D.F.H.-R., E.O.-M., Conceptualization, Data curation, Formal analysis, Methodology, Writing – original draft, Writing – review & editing, E.C.-A., M.A.V., Data curation, Formal -analysis, Writing -review & editing–, and J.F.-C, G.T.-V., Conceptualization, Data curation, Formal analysis Investigation, Methodology Supervision, Writing – original draft, Writing – review & editing.

---

**Supplementary Table 1.** Antibodies for immunohistochemistry

| Antibody | Origin | Antigen retrieval | Dilution | Origin |
| --- | --- | --- | --- | --- |
| VIP | Mouse monoclonal | Citrate buffer of pH 6 | 1:150 | Santa Cruz Biotechnology, |
| c-kit (CD117) | Rabbit monoclonal | Citrate buffer of pH 6 | 1:500 | BioSB, Santa Barbara, CA |
| GAD65 | Rabbit polyclonal | Citrate buffer of pH 6 | 1:100 | Abcam |
| PNMa | Rabbit polyclonal | EDTA buffer of pH 8 | 1:100 | Abcam |
| S100 | Mouse monoclonal | Citrate buffer of pH 6 | 1:3000 | Abcam |
| P Substance | Mouse monoclonal | Citrate buffer of pH 6 | 1:3000 | Abcam |
| REGA-3G12 | Mouse monoclonal | Citrate buffer of pH 6 | 1:100 | Rega Institute |
| REGA-2D9 | Mouse monoclonal | Citrate buffer of pH 6 | 1:100 | Rega Institute |

